# Shoulder MRI Findings in Manual Wheelchair Users with Spinal Cord Injury

**DOI:** 10.1101/2020.07.29.20164673

**Authors:** Omid Jahanian, Meegan G. Van Straaten, Brianna M. Goodwin, Ryan J. Lennon, Jonathan D. Barlow, Naveen S. Murthy, Melissa M. B. Morrow

## Abstract

**Objective:** To investigate the prevalence of rotator cuff and long head of the biceps pathologies in manual wheelchair (MWC) users with spinal cord injury (SCI).

**Design:** Cross-sectional study.

**Setting:** Academic medical center.

**Participants:** MWC users with SCI.

**Outcome Measures:** Participants’ demographic and anthropometric information, presence of shoulder pain, wheelchair user’s pain Index (WUSPI) scores, and MRI findings of shoulder pathologies including tendinopathy, tendon tears, and muscle atrophy.

**Results:** Forty-four adult MWC users with SCI participated in the study. Fifty-nine percent of the participants reported some shoulder pain. The prevalence of any tendinopathy across the rotator cuff and the long head of biceps tendon was 98%. The prevalence of tendinopathy in the supraspinatus was 86%, infraspinatus was 91%, subscapularis was 75%, and biceps was 57%. The majority of tendinopathies had mild or moderate severity. The prevalence of any tears was 68%. The prevalence of tendon tears in the supraspinatus was 48%, infraspinatus was 36%, subscapularis was 43%, and biceps was 12%. The majority of the tears were partial-thickness tears. Participants without tendon tears were significantly younger (*p* < 0.001) and had been dependent on wheelchair for significantly shorter time (*p* = 0.005) than those with tendon tears.

**Conclusion:** Mild and moderate shoulder tendinopathy and partial-thickness tendon tears were highly prevalent in MWC users with SCI. Additionally, the findings of this study suggest that strategies for monitoring shoulder pathologies in this population should not be overly reliant on patient-reported pain, but perhaps more concerned with years of wheelchair use and age.

## Introduction

Spinal cord injury (SCI) is one of the leading causes of manual wheelchair (MWC) use. The number of people with SCI in the US is estimated at 249,000 to 363,000 persons with over 17,000 new SCI cases occurring each year (1). The majority of this population is non-ambulatory and extensively dependent on their upper limbs for both mobility and activities of daily living (ADLs). This places their upper extremities at high risk of repetitive strain injuries which could lead to decreased quality of life (2). High prevalence (40% to 70%) of upper extremity pain have been reported among persons with SCI which is often chronic in nature and worsens over time (2-5). Among MWC users with SCI, the shoulder is the most common site of musculoskeletal pain with prevalence of 59% to 68% (4).

There is an overall consensus in the literature of a high prevalence of rotator cuff pathology based on imaging findings in MWC users with SCI (6-10). Rotator cuff pathology is a spectrum of disease ranging from mild tendinopathy to complete tendon tears and muscle atrophy. Using magnetic resonance imaging (MRI), Akabr et al. found that 63% of chronic MWC users (duration of MWC use > 30 yrs) with SCI had rotator cuff tears (6), and this finding was further supported by a follow-up study with a larger sample size (n=299) that reported rotator cuff tendon tears in 49% of participants (7). Similarly, across multiple studies that used MR diagnostic imaging, rotator cuff tears have been reported in approximately 60% of the participants (8, 11), and rotator cuff tendinopathy has been reported in the range of 73-100% of MWC users with SCI based on MRI or ultrasound findings (9-11). In comparison to able-bodied individuals, there are significantly higher rates of rotator cuff tears in MWC users with SCI (6, 8). The prevalence of rotator cuff tears has been reported to be four times higher in a paraplegic group than in an age and sex matched control group (63% compared with 15%, respectively) (6).

The mechanisms and risk factors by which rotator cuff pathology occurs are a combination of intrinsic and extrinsic factors. Age and diabetes are among the most commonly cited intrinsic factors (12, 13). Manual and overhead activities, and sport activities such as weight training and swimming are among the extrinsic factors which have been found as significant risk factors for rotator cuff pathologies in able-bodied populations (12-15). In addition to the aforementioned factors, MWC use is significantly associated with noticeably high prevalence of rotator cuff pathologies in individuals with SCI (7, 8).

The anatomic design of the glenohumeral joint favors manipulation and mobility at the expense of stability, demanding more of the vulnerable rotator cuff (16). In MWC users, the anatomic design of the shoulder is tested further with continuous use of the upper limbs for repetitive and weight bearing activities such as MWC propulsion, pressure relief, transfers, and other activities of daily living (17). Escobedo et al. found a significant relationship between the severity of rotator cuff tears and the duration of wheelchair use (8). However, there remains limited understanding about the specific mechanisms that worsen rotator cuff pathology and modifiable risk factors for MWC users with SCI.

In spite of high prevalence of shoulder pathologies in MWC users with SCI and its negative effects on their quality of life, there are a limited number of studies describing a detailed investigation of the prevalence of disease in all rotator cuff tendons and the associated demographic factors in this population. The risk factors that significantly contribute to the incidence of rotator cuff pathologies have been studied extensively for able-bodied individuals, with far less data on patients with SCI. Furthermore, the majority of shoulder MRI studies in MWC users with SCI were conducted between 1996-2007 using MRI with field strength of 1-1.5 Tesla (T) (6-8, 18). It has been shown that MRI field strength is a determinant for diagnostic accuracy with higher field strengths leading to significantly higher sensitivities (19, 20). The purpose of this study is to investigate and report the prevalence of rotator cuff and long head of the biceps pathologies including tendinopathy, tendon tear, and muscle atrophy in MWC users with SCI and the associated risk factors including age and duration of wheelchair use.

## Methods

### Study Participants

This study was approved by the Mayo Clinic Institutional Review Board. MWC users with traumatic or non-traumatic SCIs were identified through querying medical records and care providers of local clinics that serve individuals with SCI. Participants in the current study were originally recruited for a longitudinal study “Natural History of Shoulder Pathology in Wheelchair Users.” The MRI data from the baseline visit of the participants were used for the current study. Individuals with SCI who were using a MWC as their main mode of mobility were considered for inclusion in the study if they were between 18-70 years of age and had functional upper extremity range of motion. Functional range of motion was defined as active shoulder flexion and abduction of at least 150° and the ability of the participant to touch the opposite shoulder, the back of his/her neck and his/her low back. Participants were excluded if they self-reported having a previous diagnosis of a complete shoulder tendon tear bilaterally, which would not allow longitudinal study of that tendon, or had implanted devices that were not MRI compatible.

### Special Tests and Presence of Pain

Upon enrollment, participants attended an in-lab visit and their demographic and anthropometric information was collected, including age, sex, height, weight, date/type/level of SCI, and hand dominance. One investigator (MGVS), a licensed physical therapist, performed an abbreviated clinical shoulder examination on all participants. This examination consisted of bilateral measures of active and passive range of motion, and special tests including the Neer, Hawkins-Kennedy, empty can, and external rotation resistance tests (impingement tests), and Speed’s test (long head of biceps tendinitis) (21-24). Report of anterolateral shoulder pain during these tests was recorded as a “positive” finding, and absence of pain was “negative.” Positive findings were scored as one and negative findings were scored as zero. The scores for the impingement tests (Neer, Hawkins-Kennedy, Empty can, and Resistance ER tests) were added together and reported as a lumped positive score out of a maximum total of four.

Participants were asked if they had shoulder pain on either or both shoulders. They were also asked to indicate the location of any pain at the present time on a body diagram. Self-reported shoulder pain was measured by completing the Wheelchair User Shoulder Pain Index (WUSPI). Although the WUSPI was designed to document overall shoulder pain regardless of side, participants in this study completed two WUSPI surveys, for both the right and left shoulders. To complete the WUSPI, participants were asked to rate their shoulder pain when completing 15 tasks on a 10 cm visual analog scale between “no pain” and “worst pain ever experienced” (25). Possible total scores ranged from 0 (no pain during any activity) to 150 (worst pain ever experienced during all tasks). The validity and reliability of the WUSPI has been previously reported (26).

### Magnetic Resonance Imaging of the Shoulder

Bilateral shoulders of all participants were imaged using a standard clinical non-contrast imaging protocol using a dedicated, commercially available, shoulder coil. All participants were imaged at 3 Tesla except for two who required 1.5 T due to MRI safety protocol. The participants were positioned supine with the arm and wrist in a neutral position by their side. Axial, oblique sagittal, and oblique coronal T2 FSE FS (TR4000; TE 50); Axial and oblique coronal proton density (TR 3200–3500; TE 32-33); and oblique sagittal T1 (TR 700-900; TE min) image sequences were obtained with a matrix acquisition range of 256–384 × 256, 2 NEX, and 14 cm FOV.

The images were assessed by a board-certified, fellowship-trained, musculoskeletal radiologist with 13 years of experience (NSM) using a standardized MRI Assessment of the Shoulder (MAS) guide that was previously developed by the study authors [11]. The radiologist was masked to symptoms and participant’s characteristics. Tendinopathy was classified as mild, moderate, or severe (Fig 1A-C), (27). Tears of the supraspinatus, infraspinatus, subscapularis, and teres minor tendons were classified as partial, full, or complete, located in one of three anatomical zones: insertion, tendon, or critical zone (Fig 1D-E). The region of the tear was classified as intrasubstance, bursal, or articular. For the supraspinatus, infraspinatus, and teres minor, the portion of the tendon was defined as anterior, middle, or posterior. The subscapularis portions were defined as superior, middle, or inferior. A long head of the biceps tear was categorized as partial, split, or complete with locations defined as extra-articular, intra-articular, or bicep anchor. Rotator cuff fatty muscle degeneration was classified as mild, moderate, or advanced (28).

**Figure 1.**
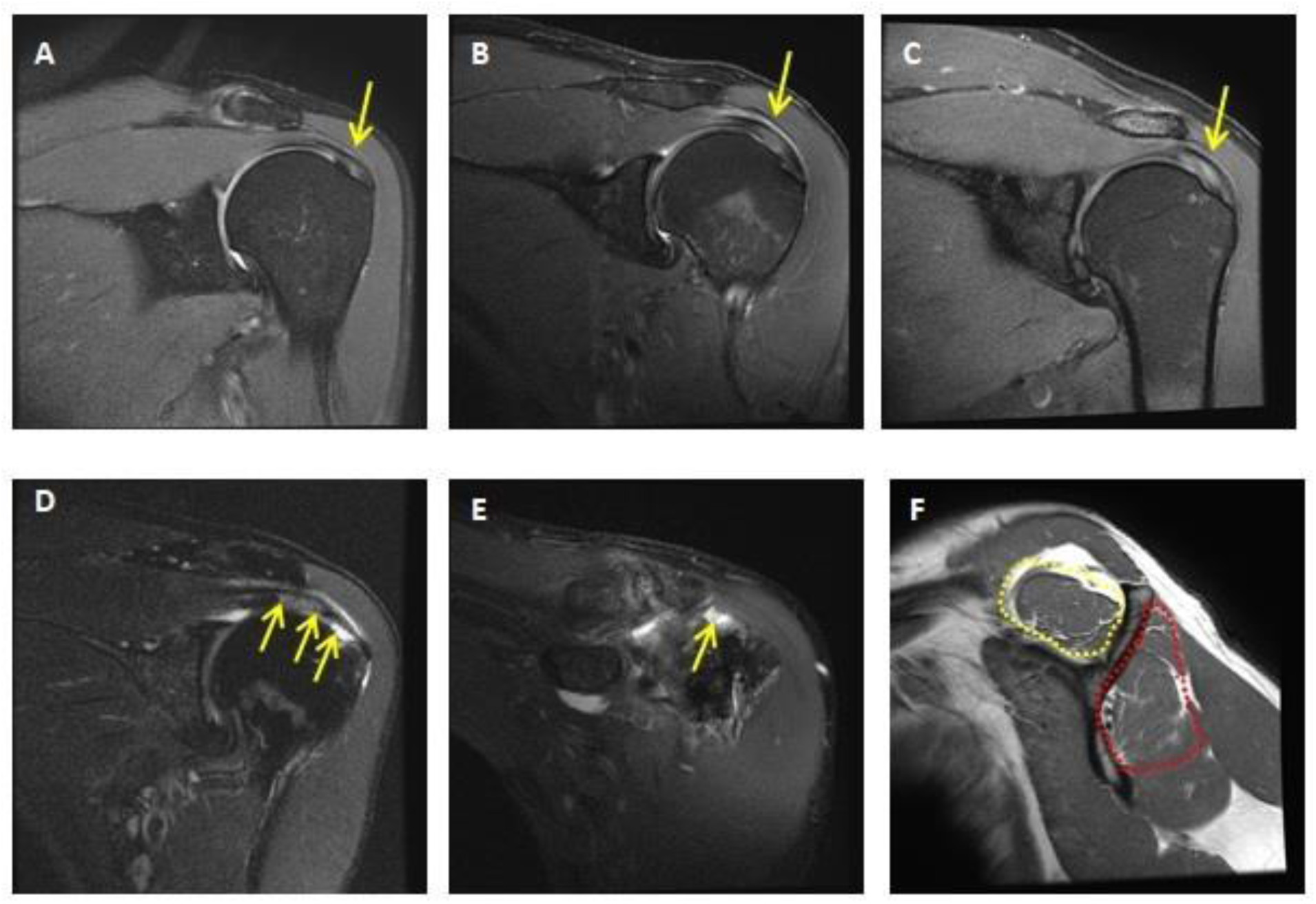
Examples of shoulder MR images illustrating tendinopathy grading, tendon tears, and fatty muscle degeneration. Mild (A), moderate (B), and severe (C) tendinopathy of the infraspinatus. (D) Partial thickness articular surface tear of the supraspinatus. (E) Full-thickness tear of the anterior supraspinatus. (F) Mild fatty muscle degeneration of the supraspinatus (yellow dotted line) and infraspinatus (red dotted line).

### Statistics

Frequencies and percentages were used to present the prevalence of shoulder pathologies including tendinopathy, tendon tear, and muscle atrophy in MWC users with SCI. Percentages were calculated based on those with available (non-missing) data. Continuous variables are summarized with mean (standard deviation) and with percentile statistics. Differences between subjects with and without any tear were compared using Student’s two sample t-test for continuous variables (age, duration of wheelchair use) and Pearson’s chi-squared test for discrete variables (sex, level of injury). The association between hand dominance and the shoulder MRI findings were tested using McNemar’s test. All significance tests are two-sided with a 0.05 Type I error rate.

## Results

Forty-four adult MWC users with SCI participated in the study. The median (IQR) participant age was 38 (35, 53) years, and 36 (82%) of the participants were men, which was representative of the SCI population in the US [1]. The median (IQR) time since injury (duration of wheelchair use) was 6.7 (1.8, 21.1) years. The mean body mass index was 25 ± 4 kg/m^2^. Among the participants there were 37 individuals with paraplegia (SCI level: T1-L1) and seven participants with tetraplegia (SCI level: C6-C8). A summary of the participant characteristics is reported in Table 1.

**Table 1.** Participant characteristics and WUSPI scores

|  |  |
| --- | --- |
| **Age , yrs**<br>Mean (SD)<br>Median (IQR)<br>Range | 42(13)<br>38 (35, 53)<br>22 - 65 |
| **Duration of wheelchair Use , yrs**<br>Mean (SD)<br>Median (IQR)<br>Range | 12.3 (12.5)<br>6.7 (1.8, 21.1)<br>0.1 – 45.3 |
| **Sex, N(%)**<br>Females<br>Males | 8 (18%)<br>36 (82%) |
| **SCI Level, N(%)**<br>C6-C8<br>T1-T8<br>T9-L1 | 7 (16%)<br>21 (48%)<br>16 (36%) |
| **SCI Lesion, N(%)**<br>Complete<br>Incomplete<br>Missing, N | 26 (62%)<br>16 (16%)<br>2 |
| **Hand Dominance, N(%)**<br>Right<br>Left | 35 (80)<br>9 (20) |
| **Height, cm**<br>Mean (SD)<br>Median (IQR)<br>Range<br>Missing, N | 179 (10)<br>179 (173 -183)<br>160 – 196<br>8 |
| **Weight, m**<br>Mean (SD)<br>Median (IQR)<br>Range<br>Missing, N | 80 (16)<br>80 (68 – 88)<br>57 – 131<br>9 |
| **Body Mass Index ,kg/m2**<br>Mean (SD)<br>Median (IQR)<br>Range<br>Missing, N | 25 (4)<br>25(22 - 27)<br>17 – 38<br>9 |
| **Cause of Injury, N(%)**<br>Motor Vehicle Accident<br>Falls<br>Sports and Recreation Injuries<br>Diseases<br>Other | 20 (45%)<br>10 (23%)<br>7 (16%)<br>3 (7%)<br>4 (9%) |
| **Non-dominant WUSPI**<br>Mean (SD)<br>Median (IQR)<br>Range<br>Missing, N | 9 (15)<br>4 (0 – 11)<br>0 – 72<br>1 |
| **Dominant WUSPI**<br>Mean (SD)<br>Median (IQR)<br>Range<br>Missing, N | 11 (17)<br>4 (0 – 15)<br>0 – 66<br>1 |

Fifty-nine percent (n = 26) of the participants reported some shoulder pain when asked if they experience shoulder pain. The prevalence of bilateral shoulder pain was 25% (n = 11), dominant side 23% (n = 10), and non-dominant side was 11% (n = 5). The average WUSPI score for dominant shoulder was 11 and for non-dominant side was 9 (Table 1). The results from the Speed’s test and the lumped scores of the impingement tests are reported in Table 2.

**Table 2.** Shoulder special test scores

|  | Score | Dominant Side | Non-dominant Side |
| --- | --- | --- | --- |
|  |  | N (%) | N (%) |
| **Speed's test** ¥ | 0 | 32 (80%) | 36 (88%) |
|  | 1 | 8 (20%) | 5 (12%) |
| **Impingement tests** ¥<br>(Neer + Hawkins-Kennedy + Empty can +<br>Resistance ER) | 0 | 27 (67%) | 29 (71%) |
|  | 1 | 3 (8%) | 7 (17%) |
|  | 2 | 4 (10%) | 3 (7%) |
|  | 3 | 4 (10%) | 1 (2.5%) |
|  | 4 | 2 (5%) | 1 (2.5%) |

¥ The data on shoulder special test scores are missing for two participants.

The shoulder MRI findings indicated that the prevalence of any tendinopathy across the rotator cuff and the long head of biceps tendon was 98% (n = 43; Figure 2). Eighty-six percent (n = 38) of the participants had supraspinatus tendinopathy. The prevalence of infraspinatus tendinopathy was 91% (n = 40). Subscapularis tendinopathy was present in 75% (n = 33) of the participants. Additionally, 57% (n = 25) of the participants had long head of biceps tendinopathy. The majority of tendinopathies had mild or moderate severity (Figure 2).

**Figure 2.**
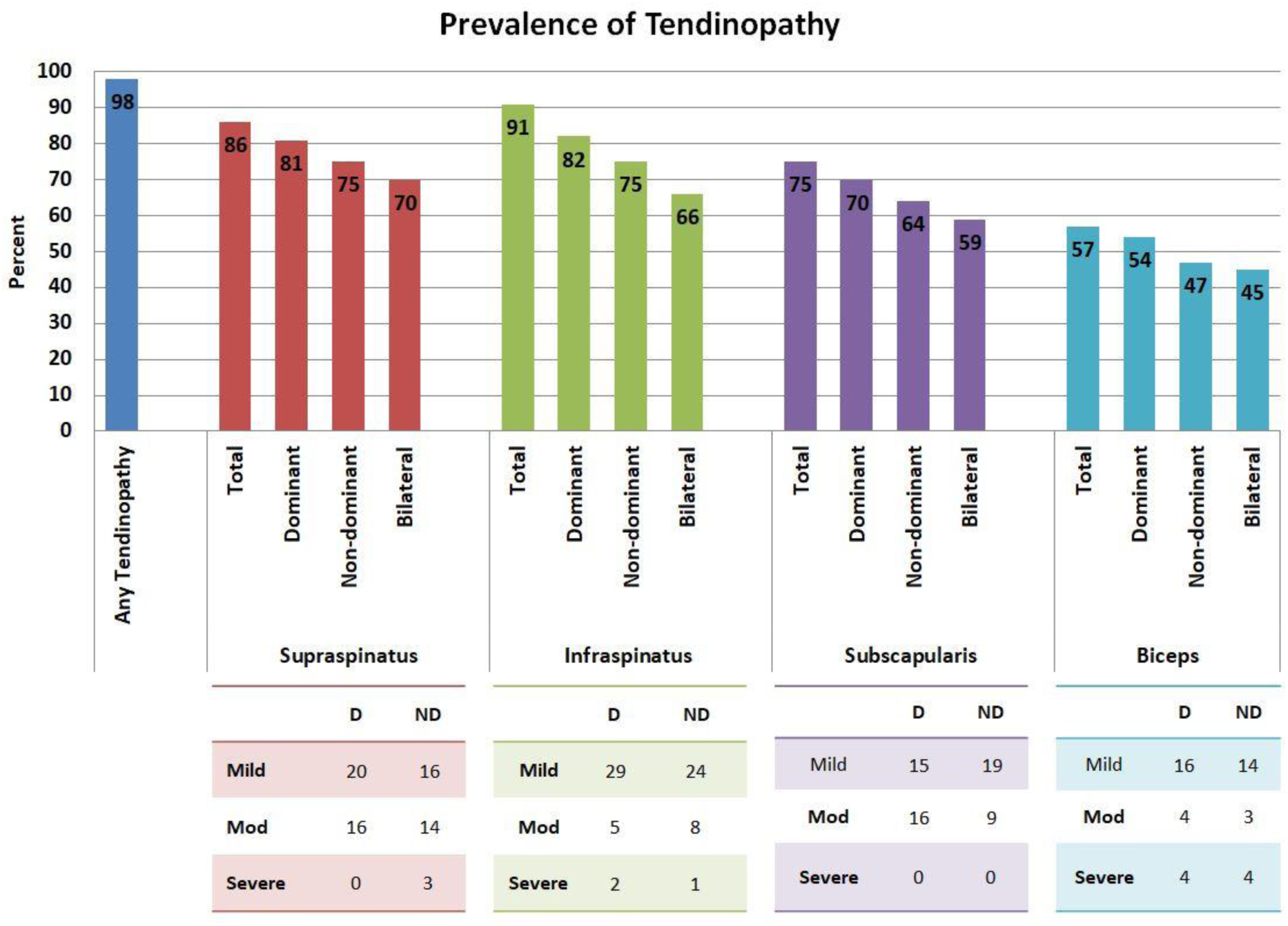
Shoulder tendinopathy in MWC users with SCI. Any tendinopathy bar indicates the percent of participants with any tendinopathy (mild, moderate, or severe) across the rotator cuff muscles and the long head of biceps tendon either at the dominant shoulder, non-dominant shoulder, or bilaterally. Total bars indicate the prevalence of any tendinopathy (mild, moderate, or severe) at the dominant shoulder, non-dominant shoulder, and bilaterally. The tables at the bottom include the number of tendons with mild, moderate (Mod) and severe tendinopathies at the dominant (D) and non-dominant (ND) shoulders.

The prevalence of any tears across the rotator cuff and the long head of biceps tendon was 68% (n = 30); all were either partial or full thickness tears and there were no participants with complete tears (Figure 3). Forty-eight percent (n = 21) had a supraspinatus tendon tear either at the dominant shoulder, non-dominant shoulder, or bilaterally. The majority of the supraspinatus tendon tears were partial-thickness tears. The prevalence of tendon tear for infraspinatus and subscapularis was 36% (n = 16) and 43% (n = 19), respectively and all were partial thickness tears. Long head of biceps tendon tears were present in 11% (n = 5) of the participants. The majority of them were partial thickness tears (Figure 3).

**Figure 3.**
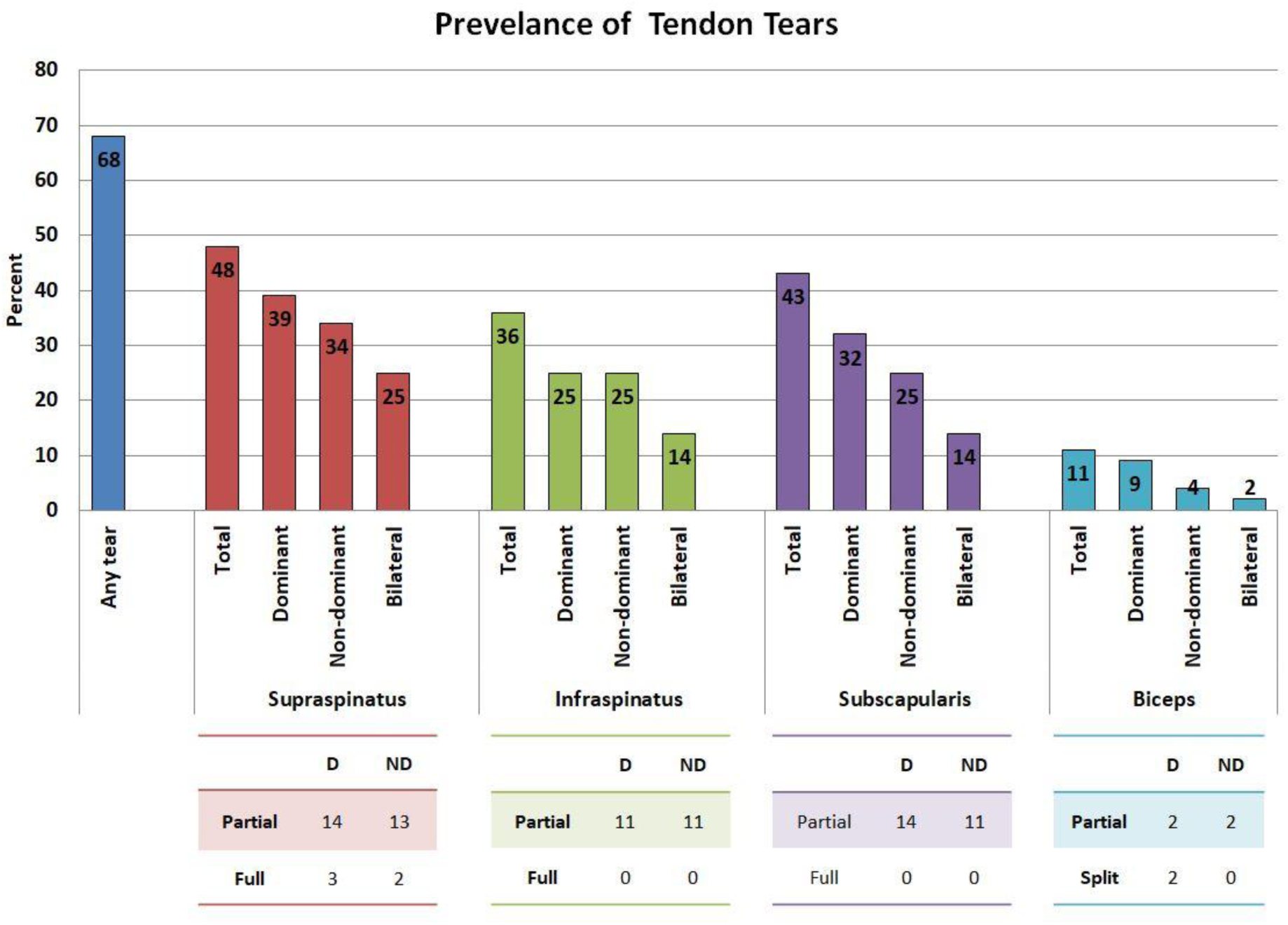
Shoulder tendon tears in MWC users with SCI. Any tear bar indicates the percent of participants with any tears (partial-thickness and full thickness) across the rotator cuff muscles and the long head of biceps tendon either at the dominant shoulder, non-dominant shoulder, or bilaterally. Total bars indicate the prevalence of any tears (partial-thickness and full thickness) at the dominant shoulder, non-dominant shoulder, and bilaterally. The tables at the bottom include the number of tendons with partial-thickness (partial) and full-thickness (Full) tears at the dominant (D) and non-dominant (ND) shoulders.

In the supraspinatus tendon, the most common locations of the partial tears were the insertion zone, bursal region, anterior portion, and middle portion of the tendon. The full tears all occurred at the insertion zone in the anterior or middle portion. In the infraspinatus tendon, the most common locations of the partial tears were the insertion zone, intra-substance region, and anterior portion. In the subscapularis tendon, the most common locations of the partial tears were the tendon zone, intra-substance region, and superior portion. The biceps anchor was the most common site of partial tears in the long head of biceps tendon. Of the two people who had a split tear of the bicep, one of the tears was in the extra-articular and the other was in the intra-articular region of the long head of the biceps tendon.

Mild fatty muscle degeneration of the rotator cuff was present in 9% (n = 4) of the participants, across the supraspinatus (7%, n = 3), infraspinatus (9%, n = 4), and subscapularis (5%, n = 2). Thirty-two percent (n = 14) of the participants had no tendon tears and in 68% (n = 30) of the participants at least one tendon tear was present. Subjects without tendon tears were significantly younger (*p* < 0.001) and had been dependent on wheelchair for significantly shorter time (*p* = 0.005) than those with tendon tears (Table 3). There was no significant association observed between sex and level of injury with the presence of a tendon tear.

**Table 3.** Associations between age and duration of wheelchair use with shoulder muscle tendon tears. IQR indicates the interquartile range (first and third quartiles).

|  | No tear (N=14) | Any tear (n=30) | *p* value |
| --- | --- | --- | --- |
| Age, yrs, median (IQR) | 32.5 (27.5, 35.8) | 51.0 (38.0, 56.0) | < 0.001 |
| Years of wheelchair use, median (IQR) | 2.0 (1.1, 5.5) | 16.1 (3.2, 24.7) | 0.005 |
| Male | 12 (86%) | 24 (80%) | 0.65 |
| Level of injury |  |  | 0.29 |
| C6-C8 | 2 (14%) | 5 (17%) |  |
| T1-T8 | 9 (64%) | 12 (40%) |  |
| T9-L1 | 3 (21%) | 13 (43%) |  |

There was no significant association between shoulder pain and the presence of a tear or tendinopathy in the related shoulder (Table 4). Patient characteristics were similar between those with and without shoulder pain. None of the shoulder MRI findings (the presence of any tear, any tendinopathy, and within specific tendons) were statistically different between the dominant and non-dominant shoulder (*p* > 0.18 for all McNemar tests).

**Table 4.**
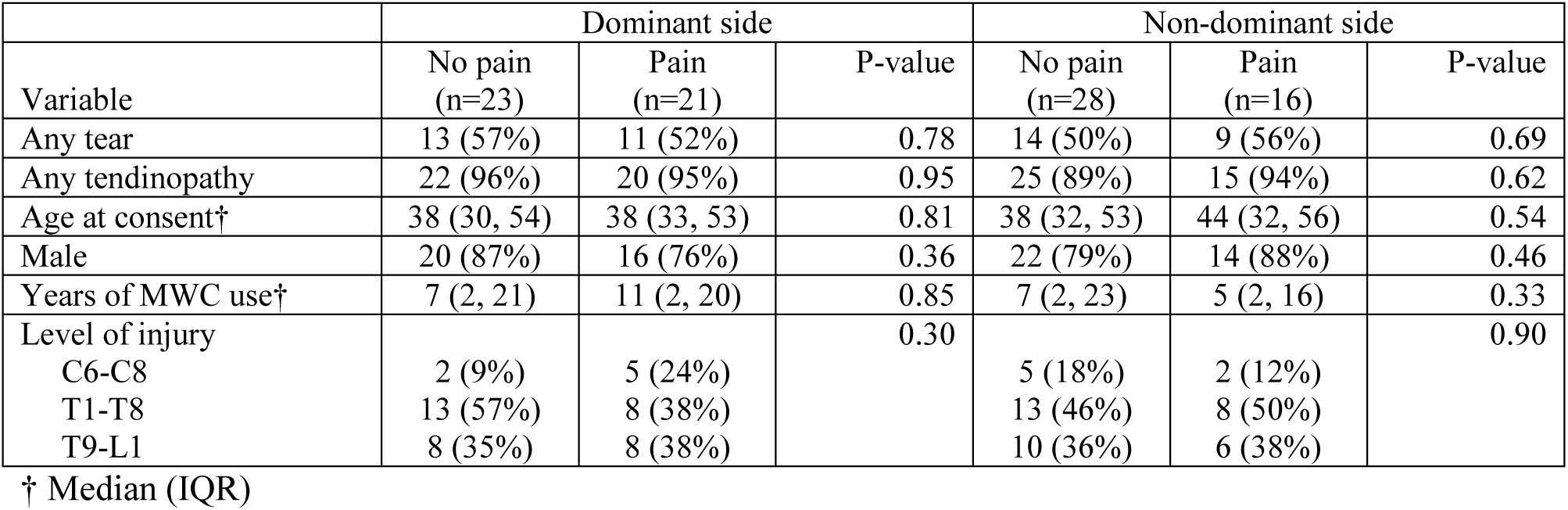
Associations between the presence of shoulder pain and shoulder pathologies, by dominance.

## Discussion

The primary goal of this study was to report detailed MRI findings of shoulder pathologies including tendinopathy, tendon tears, and muscle atrophy in adults with SCI who use manual wheelchairs. Additionally, the association between age and duration of wheelchair use with prevalence of tendon tears was tested.

In this study, 59% of the participants reported shoulder pain. The pain was considered mild in intensity as evidenced by relatively low WUSPI scores for dominant and non-dominant shoulders. These results are consistent with those previously reported in studies on shoulder pain in wheelchair users with SCI (2-4). Our results did not identify any significant associations between pain and the prevalence of shoulder pathologies. While this lack of association may differ in a cohort of individuals with higher levels of pain, a similar relationship between shoulder pain and MRI findings has been reported previously in an able-bodied cohort (29).

The findings of this study demonstrated the high prevalence of rotator cuff and long head of biceps tendinopathy and tendon tears in MWC users with SCI. The prevalence of shoulder tendinopathy was similar to what was reported in previous studies (9, 10). Since tendon pathology is a spectrum of disease which begins as tendinopathy and can develop into more debilitating tears and atrophy, it is important to describe where on the spectrum the SCI population has pathology. This knowledge may lead to better understanding about when is the best time to intervene to reduce the rate of disease advancement. In this study, infraspinatus and supraspinatus tendons had the highest prevalence of tendinopathy and the majority of the cases had mild or moderate severity; however, previous studies did not report the detailed findings regarding severity of rotator cuff tendinopathy in MWC users.

The total prevalence of rotator cuff and long head of biceps tendon tears was 68% and the majority of the tears were partial-thickness tears (92%). In the study conducted by Akbar et al. the prevalence of rotator cuff tendon tears in chronic MWC users was 63% and the majority were full-thickness tears (78%). Using MRI scanners with higher magnetic field strength (3 Tesla) than previous studies (1-1.5 Tesla) could add to the notable difference between the two studies in terms of degree of tendon tears. Greater field strength could lead to significantly higher sensitivity and better diagnosis of partial-thickness tears (19, 20). Most importantly, the participants in the current study were younger (mean age = 41.6 years) and had notably shorter mean duration of wheelchair use (12.3 years) than those who participated in Akbar’s study (average age: 52 years, mean duration of wheelchair use: 33.7 years), which is likely a large contributor to the differences in prevalence findings between the two studies.

In this study, supraspinatus (48%), subscapularis (43%), and infraspinatus (36%) had the highest prevalence of tendon tears. No biceps tendon tears were observed as an isolated finding. Escobedo et al. theorized this as an implication that long head of biceps tendon tear is a part of the spectrum of progression of rotator cuff disease which might be an indication of greater severity (8). Previous studies reported the highest rate of tendon tears for the supraspinatus (6, 8); however the prevalence of subscapularis tendon tears was notably lower in these studies in comparison with the current study. The differences in the prevalence of subscapularis tendon tears may be partially explained by the utilization of MRI machines with magnetic field strength of 3 Tesla for the majority of participants in the current study. Previous studies have demonstrated the lower accuracy of 1-1.5 Tesla MR imaging in diagnosis of partial-thickness tears than full thickness tears (30, 31). There is the potential that the studies which used 1-1.5 Tesla MR imaging systems have underestimated the prevalence of subscapularis partial tears.

As expected, the findings of this study demonstrated that age and duration of wheelchair use are both significant risk factors for shoulder tendon tears in MWC users with SCI. This is consistent with the results reported by previous shoulder MRI studies (6-8, 19). The median age and duration of wheelchair use for individuals with no tendon tears were 32.5 years and 2 years, respectively. This is significantly less than the median age (51 years) and duration of wheelchair use (16.1 years) for those with tendon tears. The significant increased risk of tendon tears with increasing age shown in this study is consistent with the knowledge regarding age-related degenerative changes such as decreased elasticity and decreased overall tensile strength in aging rotator cuff tendons (13, 32, 33). The intrinsic and extrinsic factors contributing in the incidence of rotator cuff pathologies could be amplified by overuse (34). Individuals with SCI who are dependent on MWCs, use their upper limbs for repetitive and weight bearing activities such as MWC propulsion, pressure relief, and transfers, as well as activities of daily living. This could increase the risk of rotator cuff pathologies in this population as their duration of wheelchair use increases. Additionally, the sitting position in MWC users may require frequent overhead reaching postures which could also increase the risk of sub-acromial impingement (14).

In addition to age and duration of wheelchair use, we also explored the effects of other parameters including sex, level of injury, and hand dominance. Boninger et al. reported that women with SCI who use MWC may be at higher risk of shoulder injuries than men (17). Although we did not identify an association between sex and shoulder pathology in this study, the number of women in this analysis was low (N = 8) and readers should be cautious about drawing conclusions about sex differences in this data set. Hand dominance was not associated with the prevalence of shoulder pathologies. Additionally, we did not observe a significant association between tendon pathologies and level of injury; however, larger sample sizes of each level of injury for adequate statistical power are needed to draw conclusions.

All the MR images in this study were reviewed by one musculoskeletal radiologist who was masked to symptoms and participant’s characteristics. However, the addition of multiple readers would improve the rigor of the study. The relatively small sample size is a limitation of this study which limited the interpretation of the findings regarding the contribution of the risk factors in incidence of shoulder pathologies in MWC users with SCI. The data reported were cross-sectional; therefore, it was not possible to separately analyze the effects of age and duration of wheelchair use on the incidence of rotator cuff pathologies. This will be addressed in the longitudinal study which is ongoing.

The findings of this study demonstrated the high prevalence of mild shoulder tendinopathy and partial-thickness tendon tears in a sample of relatively young MWC users with SCI. Additionally, the findings of this study suggest that strategies for monitoring shoulder pathologies in this population should not be overly reliant on patient-reported pain, but perhaps more concerned with years of wheelchair use and age. Further longitudinal investigation with a larger population of MWC users with SCI is necessary to elucidate the evolution of shoulder pathology and function due to MWC use.

## Data Availability

All data referred to in the manuscript are avialable per request.

## Conflicts of Interest

There are no conflicts of interest to disclose.

## Acknowledgements

This publication was made possible by funding from the National Institutes of Health (grant no. R01 HD84423-01), Mayo Clinic Robert D. and Patricia E. Kern Center for the Science of Health Care Delivery, and the National Center for Advancing Translational Sciences (UL1 TR002377).

## Notes

### Competing Interest Statement

The authors have declared no competing interest.

### Author Declarations

This study was approved by the Mayo Clinic Institutional Review Board.

